# WMHs AND HOMOCYSTEINE LEVELS AS CANDIDATE BIOMARKER FOR COGNITIVE IMPAIRMENT

**DOI:** 10.1101/2020.05.04.20090704

**Authors:** Vivek Misra, Ennapadam S. Krishnamoorthy, Shruti Balaji, Jayandrakumar Kuppan, K Nagarajan

## Abstract

The aim of this pilot study was to test the hypotheses that a) WMHs would be a biological marker in South Indian patients who present with either cognitive impairment, dementia or depression and b) presence of WMHs would be related to other blood biochemical markers. A significant positive correlation was present between temporal WMHs and increased homocysteine levels. A significant inverse correlation was obtained between MMSE scores and frontal caps. Association between temporal WMHs and elevated plasma homocysteine levels implies that this can serve as a biological marker for cognitive impairment and possibly for therapy.

## INTRODUCTION

Brain subcortical, periventricular and deep white matter hyperintensities (WMHs) are a frequent finding in magnetic resonance (MR) imaging of the elderly^1,2^. They are significantly associated with an increased risk of stroke, dementia, and death^3^ and with common cardiovascular risk factors such as hyperhomocysteinemia^4^ hypertension and hypercholesterolemia^5^. Recently their association with various neurobehavioral abnormalities, in particular, cognitive impairment and depression in otherwise normal elderly, has been extensively studied.

Much research provides evidence in favor of a significant inverse correlation between white matter lesions (WMLs) and cognitive decline^6^. The early stage of dementia in which cognitive impairment due to a vascular etiology does not significantly affect an individual’s daily activities has been termed “Vascular Cognitive Impairment” (VCI)^6^.

A number of studies have also highlighted the relationship between WMLs and depression. One consistent finding in MRI studies is an increased frequency and severity of subcortical WMHs in old, depressed individuals^7^. Therefore consistent with VCI, the term “Vascular Depression” has been proposed.

However it remains controversial whether the extent of WMLs per se is an independent predictor of cognitive impairment, depression, other neurobehavioral abnormality; and whether WML alone or in combination with other biomarkers is a significant predictor of outcome.

## METHODS

### Setting

Voluntary Health Services, a large community hospital with 14 primary health centers.

### Subjects

All participants aged 55 years and over, who were screened positive for cognitive impairment as part of an ongoing clinical trial of a plant based formulation. Exclusion criteria: Subjects with pre-existing medical or psychiatric condition that precluded participation; subjects on medication for cognitive enhancement, both traditional and modern (including anti-dementia drugs). 40 subjects were identified as being suitable, 29 (72.5%) agreed to participate (9 men), provided written informed consent. Approval for our study was obtained from the human research ethics committee of the Voluntary Health Services Hospital.

### MRI Scan

All consenting subjects were imaged using 1.5 Tesla, MAGNETOM Avanto Siemens Tim scanner (Siemens AG, Erlangen, Germany). After scout three dimensional T1 weighted sequences were obtained for whole brain with repetition time (TR) = 20ms; echo time (TE) = 2.1ms; bandwidth (BW) = 150 Hz; field of view (FOV) = 230*230; matrix size = 256*256; slice thickness = 5mm; flip angle 20°. The T2 weighted fluid-attenuated inversion recovery (FLAIR) sequences were obtained with TI = 2500ms; TR = 9000ms; TE = 100ms; BW 150 Hz; time acquisition (TA) = 160s; FOV= 230*230mm; matrix size = 256*256mm; slice thickness = 5mm; turbo factor = 19; flip angle 20°. MRI scans were transferred to a Windows XP workstation and analyzed using imaging suites SPM 5^8^ and MRIcro^9^. T2-weighted MR images were scored according to semi-quantitative rating scales devised by Fazekas et al^10^., modified Scheltens scale^11^ and Manolio scale^12^.

### Cognitive Measures

A brief battery of cognitive tests was administered to the subjects which include Mini Mental State Examination (MMSE), Hospital Anxiety and Depression Scales (HADS).

### Biochemical Measures

Biochemical measures include Very Low Density Lipoprotein (VLDL), Low Density Lipoprotein (LDL), High Density Lipoprotein (HDL), Thyroglobulin (Tg), Folic acid, homocysteine (Hcy), Vitamin B12 and serum globulin. Baseline investigations were also carried out which include fasting lipid analyses, coagulation studies, glucose and insulin measurements.

### Statistical Methods

The data were analyzed using SPSS (IBM v. 20, Chicago, IL, USA). Linear regression analyses were performed to determine the contribution of WMHs to the biochemical variables. Bivariate analysis using Pearson’s correlation coefficient was done to examine the association of WMH with MMSE and HADS.

## RESULTS

Table 1 details the descriptive characteristics of the sample.

**Table 1:**
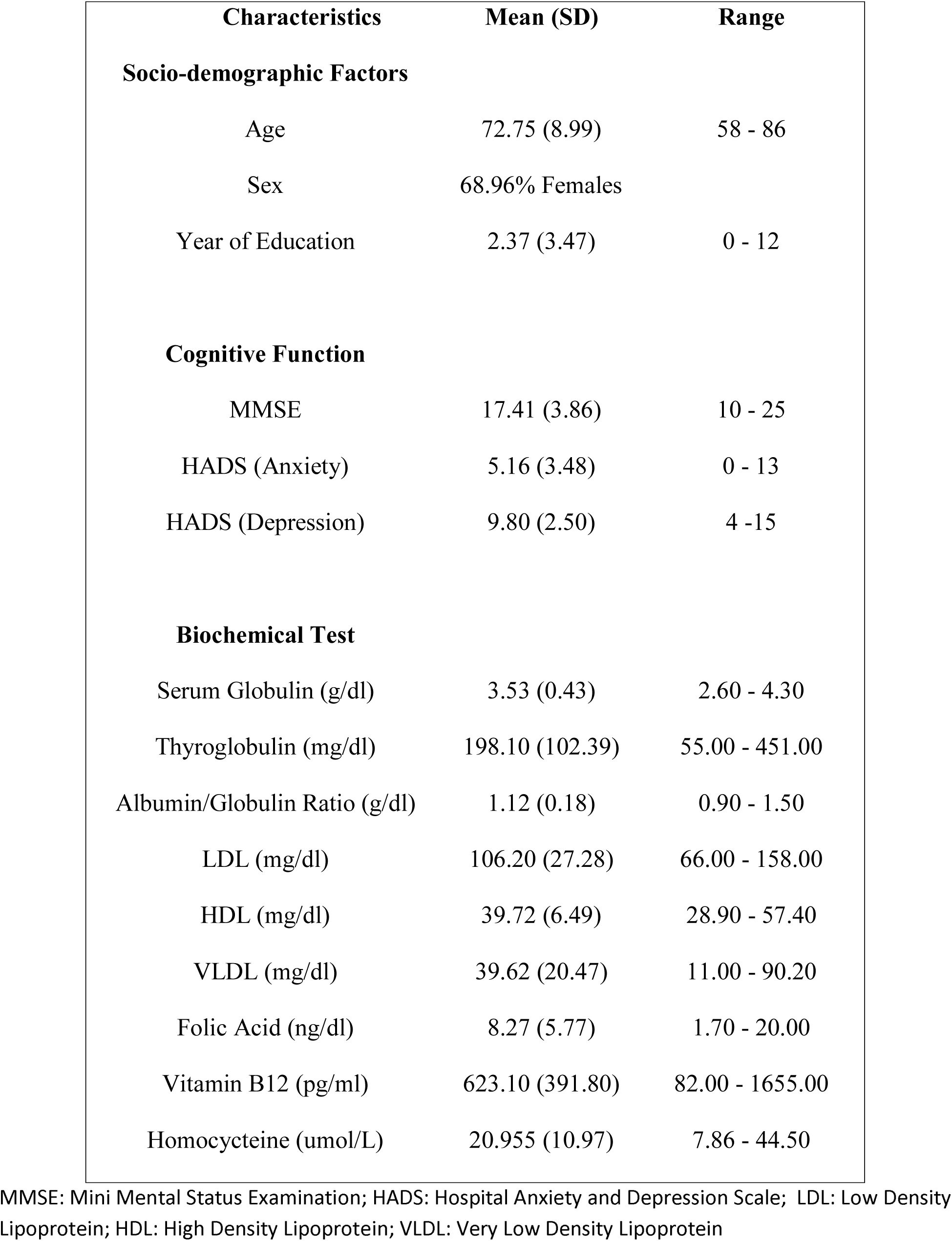
Characteristic of the sample.

Most common biochemical parameters found is abnormal values were elevated homocysteine levels and high globulin content. A significant positive correlation was observed between temporal region WMHs (WMH-T) as scored by the modified Scheltens scale and increased homocysteine (Table 2). A significant inverse correlation was noted between MMSE scores and frontal caps as scored by the modified Scheltens scale (Table 3). In bivariate analysis Caps F scores lost their association with MMSE after controlling for age and sex. It was also found that the female sex was the only significant predictor of MMSE. We did not found any significant correlations between HADS scores and WMHs.

**Table 2:** Results of regression analysis predicting biochemical variables from WMHs: unstandardized regression (B) and standardized coefficients (G) adjusted for age and sex.

| WMH Regions | Serum<br>Globulin<br>(g/dl) | Thyroglobulin<br>(mg/dl) | A/G<br>Ratio<br>(g/dl) | LDL<br>(mg/dl) | HDL<br>(mg/dl) | VLDL<br>(mg/dl) | Folic<br>Acid<br>(ng/dl) | Vitamin<br>B12<br>(pg/ml) | Homocysteine<br>(umol/L) |
| --- | --- | --- | --- | --- | --- | --- | --- | --- | --- |
| **Frontal** |  |  |  |  |  |  |  |  |  |
| B | 0.032 | 0.005 | -1.906 | -0.004 | 0.003 | 0.023 | -0.058 | 0 | 0.065 |
| $\beta$ | 0.006 | 0.201 | -0.159 | -0.048 | 0.009 | 0.201 | -0.146 | 0.064 | 0.314 |
| p Value | 0.975 | 0.397 | 0.41 | 0.84 | 0.969 | 0.397 | 0.459 | 0.74 | 0.097 |
| **Temporal** |  |  |  |  |  |  |  |  |  |
| B | -0.483 | 0 | 0.37 | 0.003 | 0.013 | 0 | -0.027 | 0 | 0.028 |
| $\beta$ | -0.255 | 0.004 | 0.086 | 0.086 | 0.089 | 0.004 | -0.186 | -0.008 | 0.38 |
| p value | 0.181 | 0.985 | 0.659 | 0.718 | 0.71 | 0.985 | 0.343 | 0.968 | **0.042** |
| **Parietal** |  |  |  |  |  |  |  |  |  |
| B | -0.043 | 0.007 | -1.739 | 0.002 | -0.028 | 0.033 | -0.04 | 0.001 | 0.056 |
| $\beta$ | -0.009 | 0.372 | -0.155 | 0.028 | -0.088 | 0.327 | -0.107 | 0.116 | 0.286 |
| p value | 0.964 | 0.159 | 0.423 | 0.908 | 0.712 | 0.159 | 0.586 | 0.548 | 0.132 |
| **Occipital** |  |  |  |  |  |  |  |  |  |
| B | -0.961 | -0.001 | 0.691 | 0.001 | 0.066 | -0.003 | 0.001 | 0 | 0.036 |
| $\beta$ | -0.201 | -0.034 | 0.063 | 0.013 | 0.23 | -0.034 | 0.002 | -0.016 | 0.191 |
| p value | 0.296 | 0.886 | 0.744 | 0.955 | 0.329 | 0.886 | 0.993 | 0.934 | 0.32 |
| **Caps Frontal** |  |  |  |  |  |  |  |  |  |
| B | -0.028 | 0.001 | -0.164 | 0.002 | -0.017 | 0.005 | -0.002 | 0 | 0.009 |
| $\beta$ | -0.023 | 0.183 | -0.059 | 0.116 | -0.205 | 0.183 | -0.018 | 0.083 | 0.186 |
| p value | 0.905 | 0.441 | 0.761 | 0.627 | 0.386 | 0.441 | 0.926 | 0.669 | 0.334 |
| **Caps Occipital** |  |  |  |  |  |  |  |  |  |
| B | 0.474 | 0.003 | -1.229 | 0.007 | 0.015 | 0.017 | 0.041 | 0.001 | 0.004 |
| $\beta$ | 0.228 | 0.397 | -0.259 | 0.231 | 0.113 | 0.397 | 0.263 | 0.358 | 0.052 |
| p value | 0.234 | 0.083 | 0.175 | 0.326 | 0.634 | 0.083 | 0.176 | 0.056 | 0.787 |
| **Bands** |  |  |  |  |  |  |  |  |  |
| B | -0.075 | 0 | -0.062 | 0.001 | -0.004 | -0.002 | 0.007 | 0 | 0.014 |
| $\beta$ | -0.048 | -0.049 | -0.017 | 0.043 | -0.031 | -0.049 | 0.06 | -0.029 | 0.222 |
| p value | 0.804 | 0.836 | 0.929 | 0.858 | 0.896 | 0.836 | 0.764 | 0.88 | 0.248 |
| **Periventricular**<br>**Body** |  |  |  |  |  |  |  |  |  |
| B | 0.402 | 0.001 | -1.246 | -0.001 | -0.004 | 0.007 | -0.025 | 0 | 0.024 |
| $\beta$ | 0.2 | 0.162 | -0.271 | -0.029 | -0.033 | 0.162 | -0.168 | -0.062 | 0.307 |
| p value | 0.298 | 0.494 | 0.155 | 0.902 | 0.889 | 0.494 | 0.394 | 0.751 | 0.105 |
| **Deep White**<br>**Matter** |  |  |  |  |  |  |  |  |  |
| B | -0.029 | 0 | -0.698 | -0.001 | 0.028 | -0.002 | -0.013 | 0 | 0.029 |
| $\beta$ | -0.013 | -0.047 | -0.137 | -0.038 | 0.191 | -0.047 | -0.074 | 0.069 | 0.326 |
| p value | 0.946 | 0.845 | 0.478 | 0.873 | 0.42 | 0.845 | 0.707 | 0.721 | 0.084 |
| **Manolio** |  |  |  |  |  |  |  |  |  |
| B | 0.237 | 0.007 | -0.897 | -0.006 | -0.021 | 0.037 | -0.037 | 0 | 0.036 |
| $\beta$ | 0.045 | 0.348 | -0.075 | -0.08 | -0.063 | 0.348 | -0.095 | -0.007 | 0.173 |
| p value | 0.816 | 0.133 | 0.699 | 0.737 | 0.793 | 0.133 | 0.632 | 0.97 | 0.371 |

\* Significant correlation at the 0.05 level (2 -- tailed).

**Table 3:** Correlation between MMSE and WMHs

| SCALE |  | DWH | PVH | Caps F | Caps O | Bands | WMH-F | WMH-P | WMH-T | WMH-O | Manol |
| --- | --- | --- | --- | --- | --- | --- | --- | --- | --- | --- | --- |
| MMSE | Pearson's Correlation (r) | -0.2 | -0.27 | **-0.37\*** | 0.03 | -0.26 | -0.25 | -0.19 | -0.22 | -0.06 | -0.25 |
|  | Sig. 2-tailed (p) | 0.29 | 0.16 | 0.05 | 0.86 | 0.17 | 0.19 | 0.33 | 0.25 | 0.75 | 0.19 |

\* Significant correlation at the 0.05 level (2 -- tailed).

## DISCUSSION

Our study shows that temporal WMHs are related to elevated homocysteine level and cognitive impairment, which is consistent with earlier studies. ^13, 14^ Subjects with elevated level of homocysteine have poor cognition when compared to subjects with normal values. Findings also suggest that, temporal lobe WMHs are more frequent in the subjects with elevated homocysteine levels, than any other brain regions.

Elevated homocysteine levels are seen to be an independent risk factor for small vessel disease, particularly ischemic leukoariosis.^15^ An increased concentration of total homocysteine in the central nervous system also inhibits the vasodilating action of nitric oxide leading to excitotoxicity and/or a decreased availability of methionine. This would in turn have an effect on neurotransmission and thus lead to a deficit in multiple cognitive abilities^15^.

To the best of our knowledge, no previous study has shown any specific correlation between WMHs in the temporal lobe and elevated plasma homocysteine levels. While this could be a chance association, it may be significant as the temporal lobe is related to various components of cognitive functioning including memory, language, speech, comprehension and auditory processing.

Earlier studies have shown that there is a positive association between WMHs and depression^16^. In particular, it has been found that late-onset depression (onset over the age of 50) is associated with signal hyperintensities in the frontal lobe and basal ganglia. However, our study showed no association between WMHs and depression, which may have been due to the small sample size or the lack of specificity of the instrument employed.

The strength of this study lies in that it is a novel one conducted on the elderly in South India. The exclusion criteria were very stringent including the exclusion of patients with uncontrolled diabetes and hypertension which could otherwise confound the outcome of our study. Since the author (VM) graded MRI scans and author (SB) performed psychological assessments and both were blinded to the findings of the other, the study has good validity. The scales that we used to grade the WMLs viz. Fazekas, modified Scheltens and Manolio scales, are all validated. The HADS is also a validated screening tool for anxiety and depression. The MRI is the predominant clinical imaging modality in patients with cognitive impairment and hence adds to the strength of the study.

In conclusion, this study has clearly demonstrated for the first time the relationship between temporal WMH and elevated homocysteine levels. Studies of larger sample of subjects powered adequately to explore further the relationships between these variables are necessary.

## Data Availability

NA

## REFERENCES

1. Leaper S, Murray A, Lemmon H, et al. Neuropsychologic correlates of brain white matter lesions depicted on MR images: 1921 Aberdeen Birth Cohort. Radiology. 2001;221(1):51–5.

2. Longstreth W, Manolio T, Arnold A, et al. Clinical correlates of white matter findings on cranial magnetic resonance imaging of 3301 elderly people. The Cardiovascular Health Study. Stroke; a journal of cerebral circulation. 1996;27(8):1274–82.

3. Debette S, Markus H. The clinical importance of white matter hyperintensities on brain magnetic resonance imaging: systematic review and meta-analysis. BMJ (Clinical research ed). 2010;341.

4. Sachdev P. Homocysteine, cerebrovascular disease and brain atrophy. Journal of the neurological sciences. 2004;226(1–2):25–9.

5. van Dijk E, Breteler M, Schmidt R, et al. The association between blood pressure, hypertension, and cerebral white matter lesions: cardiovascular determinants of dementia study. Hypertension. 2004;44(5):625–30.

6. Bowler J. Vascular cognitive impairment. Journal of neurology, neurosurgery, and psychiatry. 2005;76 Suppl 5: 44.

7. Disabato B, Sheline Y. Biological basis of late life depression. Current psychiatry reports. 2012;14(4):273–9.

8. Statistical Parametric Mapping. 5 ed. London, UK: Wellcome Trust Centre for Neuroimaging, University College of London; 2005.

9. Rorden C, Brett M. Stereotaxic display of brain lesions. Behavioural neurology. 2000;12(4):191–200.

10. Fazekas F, Chawluk J, Alavi A, Hurtig H, Zimmerman R. MR signal abnormalities at 1.5 T in Alzheimer’s dementia and normal aging. AJR American journal of roentgenology. 1987;149(2):351–6.

11. Scheltens P, Barkhof F, Leys D, et al. A semiquantative rating scale for the assessment of signal hyperintensities on magnetic resonance imaging. Journal of the neurological sciences. 1993;114(1):7–12.

12. Manolio T, Kronmal R, Burke G, et al. Magnetic resonance abnormalities and cardiovascular disease in older adults. The Cardiovascular Health Study. Stroke; a journal of cerebral circulation. 1994;25(2):318–27.

13. Artero S, Tiemeier H, Prins N, Sabatier R, Breteler M, Ritchie K. Neuroanatomical localisation and clinical correlates of white matter lesions in the elderly. Journal of neurology, neurosurgery, and psychiatry. 2004;75(9):1304–8.

14. Söderlund H, Nilsson L-G, Berger K, et al. Cerebral changes on MRI and cognitive function: the CASCADE study. Neurobiology of aging. 2006;27(1):16–23.

15. Elias M, Sullivan L, D’Agostino R, et al. Homocysteine and cognitive performance in the Framingham offspring study: age is important. American journal of epidemiology. 2005;162(7):644–53.

16. Krishnan K, Hays J, Blazer D. MRI-defined vascular depression. The American journal of psychiatry. 1997;154(4):497–501.

